# Chikungunya masquerading as dengue infection in Sri Lanka uncovered by metagenomics

**DOI:** 10.1101/2025.05.30.25328620

**Authors:** K M Ahsanul Kabir, Chathurani Sigera, Sachith Maduranga, Praveen Weeratunga, Senaka Rajapakse, Deepika Fernando, Andrew R. Lloyd, Rowena A. Bull, Chaturaka Rodrigo

## Abstract

**Introduction:** Dengue is a significant threat to human health in South and Southeast Asia where patients are treated without diagnostic confirmation during outbreaks. This approach, though cost-effective may miss important infections especially those caused by other arboviruses (e.g., Zika, Chikungunya and West Nile virus). This study aimed to diagnose missed infections mimicking dengue by using metagenomic next generation sequencing (mNGS).

**Methods and principal findings:** Total nucleic acid (DNA and RNA) was extracted and subjected to mNGS from acute infection plasma of 60 patients from a prospective cohort study in Sri Lanka in which patients with clinically suspected dengue fever were recruited but were later confirmed as dengue-negative by NS1 antigen testing and by dengue-specific reverse transcription and polymerase chain reaction (RT-PCR) analysis.

mNGS data revealed missed chikungunya and dengue infections in five patients each, and a possible bacterial infection by *Klebsiella pneumoniae* in another patient. It was not possible to differentiate chikungunya infections from dengue infections based on clinical features or routine non-diagnostic laboratory tests conducted in early infection (e.g., full blood count, C-reactive protein level). Phylogenetic analysis showed that the chikungunya sequences from this study were closely related to those sequenced from Maldives, Malaysia, India and Singapore between 2015 – 2019.

**Conclusions:** Chikungunya infection may masquerade as dengue especially in low- and middle-income countries where dengue is treated based on clinical suspicion only - without confirmatory testing. As both infections are likely prevalent worldwide, but the complications and natural history of chikungunya and dengue infections are quite different, the addition of cheap and accessible diagnostics for both infections should be pursued in endemic countries.

**Author summary:** Dengue infection is often treated in low- and middle-income countries based on clinical suspicion only without confirming the diagnosis because of the costs of confirmatory testing. However, the clinical presentation of dengue can be mimicked by other arboviral infections like Chikungunya and Zika and these infections that need to be managed differently may be missed during dengue outbreaks. In this pilot exploratory study, we diagnosed 5 chikungunya infections as the sole acute viral infection in 60 Sri Lankan patients who were treated as for dengue during their acute febrile illness but without laboratory confirmation of dengue infection, using metagenomic next generation sequencing. Acute chikungunya infections are hardly ever reported or tested in Sri Lankan febrile patients because of its presumed rarity. Detection of multiple missed chikungunya infections in this small sample suggests the need for larger studies to evaluate the true burden of Chikungunya and other missed infections mimicking dengue.

## Introduction

Acute undifferentiated fevers (AUF) of likely viral origin are a common presentation in primary care [1]. They are typically referred to as “influenza like illnesses”, or specifically labelled as a presumptively diagnosed common viral infection such as influenza, but not always with laboratory confirmation [2, 3]. AUF are mostly self-limiting [4], and so diagnostic laboratory testing may be unwarranted. However, clinical decision-making will inevitably be aided by a precise microbiological diagnosis. Nevertheless, given the many diagnostic possibilities, the costs and logistics of microbiological testing, a confirmed diagnosis is often not pursued and this is particularly so in low- to middle-income countries (LMIC) where the available health budget is limited [2, 5]. A systematic review of publications reporting on 80,554 patients with acute fever in South and Southeast Asia between 1998 – 2019, found that 65% of them were undiagnosed [5]. Even in high-income countries like Australia, more than 50% of patients with AUF remained undiagnosed [6, 7]. In Europe, a prospective multicentre study of 765 febrile returning travellers over a 2-year period designated 60% of recruits as having AUF [8]. Of them, 24% and 21% were eventually confirmed to have an arboviral (dengue, Zika, chikungunya) and malaria infections, respectively.

Sri Lanka is a tropical island nation in the Indian Ocean with a population of 23 million. Being an LMIC, it has a high burden of AUF. Most AUFs fitting a “viral phenotype” based on symptoms and ancillary laboratory tests (e.g., full blood count) are presumed to be dengue, influenza or COVID (or other respiratory viral infections). Differentiating these from acute bacterial infections (e.g., leptospirosis, typhus) mostly depends on clinical expertise without confirmatory testing. Malaria is not considered without a recent travel history as there is no local transmission.

Previously, a cost analysis of 877 Sri Lankan patients with clinically suspected dengue fever showed that treating all of them for dengue without diagnostic confirmation was the most cost-effective way even though only 60% of them were retrospectively confirmed to have dengue [2]. This assumes that the remaining 40% had a self-limiting infection without the risk of complications or long-term sequelae. However, this is not true for infections like Japanese encephalitis and West Nile viruses that have been shown to circulate in Sri Lanka and can mimic dengue in the acute phase while causing serious complications like encephalitis [9]. An upfront diagnosis in such instances could lead to cost savings and better outcomes.

Clinical judgement is typically biased towards prevalent infections and new, uncommon or re-emerging pathogens mimicking a common infection are often missed with serious health and economic consequences for both patients and the community (e.g., tick borne diseases or Q fever being treated as influenza or COVID-19) [10, 11]. Furthermore, unavailability of specific diagnostic tests, prohibitive costs and healthcare worker attitudes (e.g., hesitancy to test on the presumption that some infections are absent or not common), also lead to misdiagnosis of infections and particularly those from non-dengue arboviruses in areas where dengue overshadows other virus infections like Zika and Chikungunya[12]. Long-term sequelae of infections like Zika, chikungunya, and Q fever, which typically present as an AUF in the acute phase, cannot be appropriately managed unless the acute infection is correctly diagnosed [13]. At the community level, the negative impact of AUFs can be felt as outbreaks that could have been better contained, increases in disability adjusted life years lost, and under-appreciation of the infectious diseases burden leading to misguided healthcare budgeting for preventive measures (e.g., vaccination).

Clinical metagenomic next generation sequencing (mNGS) which involves extracting all pathogen related genomic material (RNA and DNA) to characterise the microbiome of a clinical sample with high throughput sequencing, is becoming increasingly popular in diagnostics [14]. Given the specific challenges associated with mNGS (e.g., cost, contamination, difficulties in interpreting results), its’ use is mostly restricted to specific clinical situations like difficult to diagnose patients in an intensive care unit. However, it can also be a powerful tool to identify unusual, new or emerging pathogens in endemic circulation by screening non-critically ill patients with a specific clinical syndrome. This study aimed to use mNGS on plasma samples sourced from a Sri Lankan cohort of in-hospital patients with an acute febrile illness clinically resembling dengue fever but remained undiagnosed after testing negative for dengue.

## Methods

### Clinical samples

The samples were obtained from the Colombo Dengue Study (CDS), which is a single centre prospective cohort study recruiting clinically suspected adult dengue patients presenting within the first 96 hours of fever to the National Hospital of Sri Lanka [15]. The work described in this paper was limited to samples collected prior to the pandemic (2017 – 2020). The patients are followed up daily during the hospital admission to record the clinical outcomes, including plasma leakage and severe dengue [16]. The design of the cohort, recruitment strategy and findings from other CDS projects have been published previously [2, 15, 17, 18]. All patients recruited to CDS undergo two diagnostic tests: an NS1 dengue antigen test (a bedside immunochromatography test) and an RT-PCR test. The latter test is done retrospectively via batch processing for logistical purposes. Thus, the treating physicians are unaware of the final diagnosis when the patient is admitted to the hospital, given the relatively low sensitivity of the NS1 test [15]. Hence, all clinically suspected patients are managed as dengue fever. Neither NS1 antigen nor RT-PCR testing is regularly available for patients treated in the public healthcare system of Sri Lanka due to their cost [19]. If an enrolled patient was suspected of having a bacterial infection (e.g., leptospirosis) and were treated as such during their hospital stay, or if an alternative diagnostic test including blood culture became positive, they were excluded from CDS. There were 345 patients (39%) from Phase 1 recruitment in whom both NS1 and RT-PCR tests were negative for dengue, and so they were considered likely to have had an alternative infection or a non-infective febrile illness (termed here non-dengue fever patients or NDF).

### Nucleic acid extraction

Consecutive NDF cases (n=60) with available stored plasma samples were selected for this study. Total nucleic acids (TNA) were manually extracted from plasma samples using the MagMAX Viral/Pathogen Nucleic Acid Isolation Kit (Catalog No. A42352, Thermo Fisher Scientific, USA), following the manufacturer’s instructions using a starting volume of 200 µL of plasma. Briefly, plasma was mixed with 10 μL of Proteinase K, 530 µL of binding solution, 20 µL of magnetic beads, and incubated at 65°C for 5 minutes, and shaken at 1,050 rpm for 5 minutes. The beads were separated with magnets (10 minutes, supernatant discarded), washed sequentially with 1 mL wash buffer and ethanol (1 mL and 500 µL of 80% ethanol), air-dried for 5 minutes before adding 60 µL of elution solution, followed by incubation at 65°C for 10 minutes. The eluted TNA were separated from the beads using a magnet. The TNA purity was assessed with NanoDrop (Thermo Fisher Scientific) and the elution was split into two aliquots for DNA and RNA library preparation. Plasma samples known to be infected with dengue (n=2), Hepatitis C (n=2), Hepatitis B (n=1) and culture supernatant containing cytomegalovirus were used as controls.

### Library preparation and sequencing

For DNA library preparation, the Illumina DNA Prep kit (Catalog No. 20060059, Illumina, Inc., USA), which employs bead-bound transposons to simultaneously fragment genomic DNA (gDNA) and attach sequencing primers was used. This process produces DNA fragments with a uniform size of approximately 300 base pairs, followed by PCR amplification to enrich the library. The fragment size distributions of libraries were assessed using an Agilent 2100 Bioanalyzer and DNA amount was quantified with the Quant-iT™ PicoGreen™ assay. All the above steps were done at room temperature.

For RNA library preparation, the SMARTer Universal Low Input RNA Kit (Catalog number 634940, Takara Bio, USA) was used to convert RNA into double-stranded cDNA (dscDNA). Following the manufacturer’s instructions, 10 μL of TNA was mixed with the 3’ SMART N6 CDS Primer II A to synthesize the first cDNA strand using SMARTScribe Reverse Transcriptase and the SMARTer II A Oligonucleotide at 42°C for 90 minutes. The resulting first-strand cDNA was purified using SPRI AMPure magnetic Beads. The purified first-strand cDNA, still bound to the SPRI beads, was directly used for PCR amplification with Advantage 2 Polymerase and PCR Primer IIA to produce double-stranded cDNA. This amplified cDNA library was further purified, during which the beads were washed with 80% ethanol and eluted in a purification buffer. The amplified and purified dscDNA was digested with the restriction enzyme *RsaI* to excise the SMART adapter sequences and purified again with SPRI beads as before.

The DNA and RNA libraries were sequenced on the Illumina NovaSeq 6000 with a 2 x 150 bp paired-end sequencing platform (S4 flow cell) to generate an average of 120-150 million reads (each 150 – 300 bp) per sample.

### Data analysis

Sequencing data was analyzed using the open-source Illumina mNGS Pipeline v8.3 available on the CZID platform (https://czid.org). De-multiplexed reads were uploaded to CZID in FASTQ format. The pipeline checked data for quality control using fastp v 0.23.4 [20]. Human reads were removed using aligners Bowtie2 v 2.5.4 [21] and Hisat2 v 2.2.1 [22], and duplicate reads were eliminated by CZID-dedup v 0.1.2 [23] . The remaining reads were aligned to the NCBI nucleotide (NT) database using Minimap2 v 2.28 [24] and the non-redundant (NR) protein database using Diamond v 2.1.9 [25]. Short reads were also *de novo* assembled into contigs using SPADES v 4.0.0 [26], which were then queried against the NT and NR databases using GSNAPL [27] and RAPSearch2 [28], respectively. Finally, the pipeline generated a taxonomic report based on pathogen-specific reads per million total reads (rPM), providing a detailed profile of the microbial taxa present at the genus level. All scripts and user instructions for the CZID workflows are available at https://github.com/chanzuckerberg/czid-workflows.

If the same pathogen was confirmed in at least 5 samples, we compared the symptoms, signs, and haematological and biochemical parameters recorded in the first 96 hours of fever against those of confirmed dengue patients in a 1:2 case control design matched for age and sex using Mann-Whitney U Test (for continuous variables) and chi-square test (for categorical variables), adjusted for multiple comparisons with Bonferroni correction. A sample size calculation was not done as this was only an exploratory study.

### Confirmation of missed infections identified by mNGS

Assuming other arboviral infections (Zika and Chikungunya) to be the most common missed infections mimicking dengue, we employed the TaqMan Arbovirus Triplex Kit (Catalog Number A31747) which detects Zika, chikungunya and dengue infections to validate any positive mNGS findings for these viruses according to manufacturer’s instructions. The qPCR assays were conducted using the QuantStudio 7 Real-Time PCR System (Applied Biosystems) under the following thermal cycling conditions: reverse transcription at 50°C for 20 minutes to synthesize cDNA from RNA, initial activation at 95°C for 2 minutes, followed by 40 cycles of amplification consisting of denaturation at 95°C for 15 seconds and annealing/extension at 60°C for 1 minute. Fluorescence data were collected at each cycle, and threshold cycle (Ct) values were calculated using QuantStudio software. Each run included no-template controls (NTC) to confirm the absence of contamination.

### Phylogenetic analysis

For non-dengue RNA virus infections identified, a phylogenetic analysis was performed to compare the sequence similarity to other contemporary circulating genomes of the same pathogen globally if a consensus sequence covering at least 40% of the reference genome could be recovered from mNGS reads. Complete viral genomes for comparison were downloaded from the Bacterial and Viral Bioinformatics Resource Center (https://www.bv-brc.org/), and these were aligned with consensus sequences generated in this study using multiple sequence comparison using log-expectation (MUSCLE) algorithm [29] implemented in Geneious Prime (2022.1.1, Biomatters, New Zealand). The alignment was manually edited to remove gaps in the mNGS generated consensus sequences (entire columns of genome positions were deleted). The remaining positions were concatenated and realigned, and then used to build a maximum likelihood consensus phylogenetic tree with 100 bootstrap replicates using RaXML (Randomized Axelerated Maximum Likelihood) [30]. The consensus tree was visualized by collapsing nodes with a bootstrap support of less than 70%.

### Ethics approval

The research was approved by the Human Research Advisory Panel (Biomedical) at the University of New South Wales (HC220320). The CDS and its sample collection and storage processes were approved by the Ethics Review Committee of the University of Colombo, Sri Lanka (EC-17-080). All patients provided written informed consent for data and sample collection for research. Patient recruitment for samples described in this study started on 01/10/2017 and ended on 10/07/2018. The data were accessed for research purposes between 01/10/2017 and 01/11/2024. Deidentified clinical data were transferred to UNSW for analysis as approved by the Ethics Review Committee of the University of Colombo, Sri Lanka (EC-17-080) and stored in encrypted UNSW Onedrive, only accessible by the researchers. The sequence data generated were also securely transferred and stored in UNSW Onedrive.

## Results

### Pathogen characterisation by mNGS

Of the 345 NDF patients recruited to CDS during the pre-COVID phase, samples from 191 patients were available to be tested. Of these the first 60 samples collected were (60/191, 31%) to be tested on the mNGS workflow described above alongside the 6 control samples. The total sequencing data generated by mNGS for this study was 790.15 GB. Each sample produced 13.17 GB of data, with an average read count of 115 million.

In the analysis of RNA virus sequence data, filters were applied in the CZID workflow to retain reads from known pathogens and samples that had at least 100 pathogen specific reads (an arbitrary, conservative cut-off). With these filters, all infections in the control samples plus 6 likely chikungunya virus infections (read count range: 114 – 60143, median: 2197) and another 5 dengue virus infections (read count range: 460 – 1125971, median: 2754), were detected. When mNGS reads were mapped across corresponding pathogen reference genomes in all samples except one chikungunya infected sample, a consensus sequence covering a minimum of 1000 nucleotides were recovered. The triplex arboviral qPCR independently confirmed 5 chikungunya infections (but not the sixth sample mentioned above), and the 5 dengue infections. The number of pathogenic virus specific reads reported in each additional sample, when the above conservative filters were removed, are detailed in S1 and S2 Tables. However, given the low read counts and limited coverage across the corresponding reference genomes, these are unlikely to be true infections.

For bacterial (DNA) data analysis, samples with likely or probable acute viral infections (n=11) were excluded as the clinical picture was most consistent with a viral infection. Of the remaining samples (n=49), only those with genomic reads from a known pathogen with an abundance > 100 pathogen-specific reads per million of total reads, aligning throughout the length of the reference genome, were retained. Of these samples where only a single pathogen met all the above criteria (notionally equivalent to a positive blood culture with a single bacterial growth) was assigned as a likely infection. Such stringent criteria were necessary given the plethora of commensal and opportunistic pathogen DNA recovered due to likely contamination during sample collection and processing. On this basis, five samples with positive hits of *Pseudomonas putida* (n-2), *Aeromonas caviae* (n-2), and *Klebsiella pneumoniae* (n-1) were identified. Given that all 5 patients had no history of an immunocompromising illness (where they would be susceptible to *Pseudomonas* or *Aeromonas* infections), and as all recovered without antibiotics, only the *Klebsiella* infection was considered as a possible differential diagnosis for the presenting AUF.

### Characteristics of confirmed chikungunya infections

The sixty samples were collected between November 2017 and June 2018, and the five confirmed chikungunya infections were recruited in January (1), April (1), May (1) and June (2) of 2018. When the clinical records of the five confirmed Chikungunya infections were compared with those of confirmed dengue infections in a 1:2 case control design, matched for age and gender, no statistically significant differences were identified in: the frequency of symptoms or signs present on admission, the duration of the hospital stay, or in the haematological or biochemical parameters recorded in the first 96 hours of fever, except for the haematocrit value being significantly higher in dengue patients (Mann Whitney U test, p-value = 0.003, Table S3).

The chikungunya genomes recovered from the five confirmed infections were phylogenetically compared with contemporary circulating chikungunya sequences as mentioned above. In the BV-BRC database, 933 complete chikungunya genomes sequenced from human hosts from 57 countries were available on 27^th^ August 2024. From this, one representative sequence per calendar year, per country, was selected if they were isolated on or after 2015 (n = 44), except for Sri Lankan sequences for which there was no time limit. These sequences were aligned with 5 sequences generated in this study, and the alignment was manually edited (see Methods) to generate a concatenated alignment of 4815 nucleotide positions (41% coverage across the genome) after removing gaps. The 5 sequences from this study clustered together and were embedded within a larger cluster of Asian sequences, which stood apart from two clusters of American (North, Central, South American and Caribbean) sequences. The phylogenetically closest sequences were from Malaysia (2017), Singapore (2015), India (2019) and Maldives (2019). All previous Sri Lankan sequences (isolated between 2006 – 2008) were more phylogenetically distant (Fig 1).

**Fig 1.**
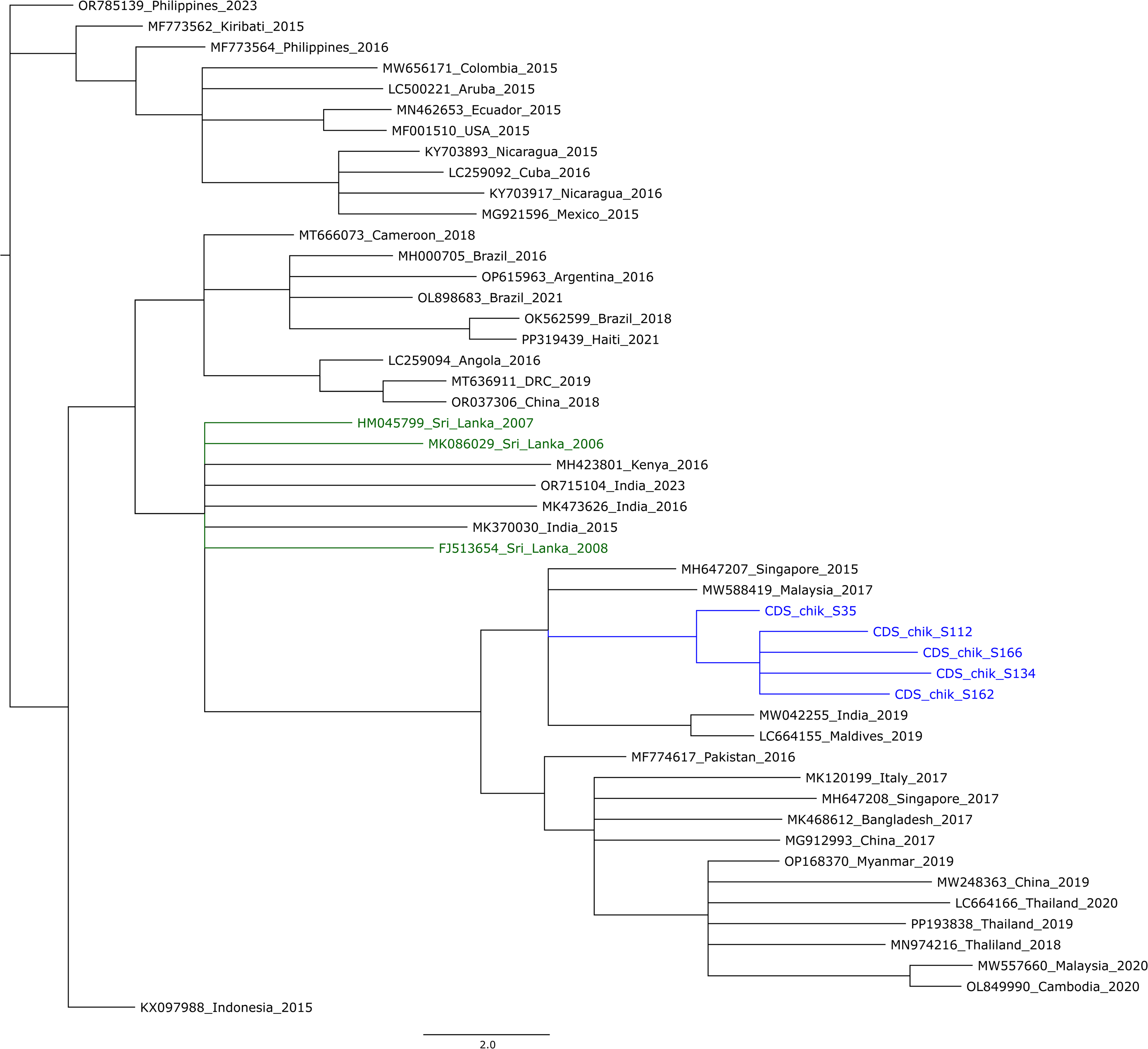
The maximum likelihood phylogenetic tree of contemporary circulating chikungunya sequences isolated from around the world. These sequences were selected from 933 complete chikungunya genomes available in the BV-BRC sequenced from human hosts from 57 countries. One representative sequence per calendar year, per country, was selected if they were isolated on or after 2015 (n = 44), except for Sri Lankan sequences for which there was no time limit. The sequences from the current study are in blue and previous sequences isolated from Sri Lanka are in green. Each tip of the tree is named as follows; Genbank accession number_Country_Year of isolation. The tree nodes collapsed to show only those with bootstrap support > 70%.

## Discussion

This exploratory mNGS analysis of 60 plasma samples from Sri Lankan patients with AUF clinically resembling dengue fever found 5 missed chikungunya infections and another five missed dengue infections. An additional sample potentially had *Klebsiella* infection, while the remainder remained undiagnosed.

The most significant finding in this study was the presence of missed chikungunya infections passing off as dengue. Clinically, it is difficult to differentiate these two infections as shown in the case control analysis where the only discerning factor was a statistically significant increase in haematocrit in dengue patients, which does not help to differentiate chikungunya cases. Chikungunya is increasingly diagnosed in dengue-endemic countries (e.g., Brazil, Cambodia, Vietnam, Bangladesh) either by mNGS [31–33] or non-mNGS[34, 35] methods. In one of the largest mNGS based studies from Cambodia, which screened 489 AUF patients, identified 138 cases of dengue infection, 13 rickettsial infections, 10 chikungunya infections and 6 malaria infections. No pathogen was detected in most cases (293 samples). Recognising the significance of missed chikungunya cases, it was subsequently added to the Cambodian national health surveillance, which identified a major outbreak across multiple provinces that exceeded 6000 cases by 2020[36].

Chikungunya is not routinely considered as a differential diagnosis for patients with AUF in Sri Lanka. While the present study cannot estimate the incidence of missed infections, finding 5 chikungunya infections within 60 tested samples, spread across several months of the first half of 2018, makes it worthwhile to investigate further if Chikungunya is endemically circulating in Sri Lanka. In fact, serological evidence for chikungunya and dengue co-circulation has been reported from Sri Lanka (or Ceylon) as far back as 1966 [37–39]. Since then, there was a paucity of published literature until 2005, when a chikungunya epidemic in La Reunion, a French overseas territory in the Indian Ocean, re-ignited interest in the topic [40]. All available Chikungunya viral sequences in BV-BRC from Sri Lanka were sequenced in studies conducted between 2006-2008 [41, 42]. Again, the interest in the infection waned and was overshadowed by the repeated dengue outbreaks until a sero-surveillance study published in 2023 identified anti-chikungunya IgG in 56 patients out of 149 tested (37.5%), indicating common prior exposure to infection [43].

Subsequently acute chikungunya infections were also confirmed independently by others in two districts of Sri Lanka in samples collected between 2017 - 2019 while this manuscript was in peer-review[44]. Compared to dengue, chikungunya is less likely to be life-threatening in the acute stage, but chikungunya related deaths have been reported during outbreaks in South Asia [45]. More commonly, in terms of morbidity, chikungunya infections can lead to chronic disabling arthritis or arthralgia [46]. If chikungunya cases are misdiagnosed as dengue, and because dengue patients are not typically followed up after recovery, patients who need long-term follow-up may not receive appropriate care.

Identifying true bacterial infections from mNGS data was more difficult given the presence of commensal and opportunistic pathogen reads alongside known pathogens probably due to contamination in a hospital setting. Furthermore, the clinical picture and routine blood investigation results of patients enrolled into CDS fit more with an acute viral fever (absence of neutrophilia) requiring more stringent filters to detect a likely bacterial infection. Of the known pathogens identified as fitting these criteria, Pseudomonas and Aeromonas infections are not typical of immunocompetent patients living in the community, and only the Klebsiella infection, which is known to cause community-acquired pneumonia, was taken as a possible explanation for the AUF.

The cost of mNGS in this study was approximately 600 AUD per sample, which rules it out for routine screening of AUF even in high income countries. Furthermore, there is no consensus on the criteria to diagnose an infection by stand-alone mNGS, and any suspected infection in a hospital patient would require validation with another accredited diagnostic test, which would add on to the cost. However, in many hospitals in high income countries, mNGS is becoming established as a backup option for ‘difficult to diagnose’ patients (e.g., immunocompromised, or in intensive care units) where an infectious cause is suspected but pathogen-specific diagnostic tests repeatedly return negative results [47, 48]. On the other hand, as shown here and by others [36], mNGS is useful to screen a limited number of samples to identify new, emerging or unusual pathogens to trigger public health surveillance with low-cost pathogen specific tests.

This study has several limitations. Firstly, this exploratory study was not based on a sample size calculation to estimate the incidence of missed chikungunya or any other infection. Furthermore, this cohort did not recruit patients by specifically querying the clinical features of chikungunya and instead found these missed infections in clinically suspected dengue patients. If chikungunya specific symptoms were queried (e.g., prominent arthralgia) it is plausible that more patients would have been found. Secondly, the sampling was from a single hospital located in Colombo city. However, this is the most populous city of Sri Lanka, and 10% of the country’s population lives within the city and the broader Colombo district and therefore still suggests a significant public health threat that must be further investigated. On top of that, Sri Lanka is an island of 65,000 sqKm where driving across from north to south or east to west can be done in less than half a day. As mentioned before Chikungunya infections have been independently confirmed in two other districts from samples collected in a similar timeframe as ours (2017 – 2019). Finally, in many samples an infection was not confirmed even after mNGS. The likely possibilities are, dengue or other arboviral infections where viraemic phase had ended at the time of sampling; bacterial or viral infections where bacteraemia or viraemia is not prominent (e.g., influenza); or fevers of non-infectious origin. This cohort recruited patients meeting the clinical case definition of dengue and all were managed as dengue during the hospital stay which meant blood cultures were not performed (as it is not required unless an alternative diagnosis is suspected). This prevented an opportunity to further validate mNGS bacterial hits against an independent confirmatory test to filter out the “background noise” of contamination.

## Conclusion

This study uncovered chikungunya infections masquerading as dengue infections using mNGS. Though the clinical symptoms of acute infection are similar in both infections, the post-acute sequelae with chikungunya are not appropriately managed when the diagnosis is missed. Other arboviral infections are typically overshadowed by the prominence of dengue infection in South Asia. Based on this finding, the next step should be the estimation of the community prevalence of chikungunya infections by screening AUF patients with a chikungunya specific test. This study shows the value of mNGS as an exploratory tool to detect novel, unusual, or re-emerging pathogens in people with AUF. Despite the high costs of initial testing, detection of missed infections can be cost saving in the long run.

## Data Availability

The mNGS raw data of this project is publicly available under the GenBank project Accession number: PRJNA1243565.

https://www.ncbi.nlm.nih.gov/bioproject/PRJNA1243565/

## Supporting information

**S1 Table. Viral pathogen reads uncovered with mNGS (only known pathogens).**

**S2 Table. Pathogenic bacterial reads uncovered with mNGS (only known pathogens).**

**S3 Table. An age and gender matched 1:2 comparison between confirmed chikungunya and dengue cases.**

## Acknowledgements

None

## Author contributions

Conceptualization: CR, experiments: AK, clinical data and samples: CS, PW, DF, SR, mNGS data analysis: AK, CR, phylogenetics data analysis: CR, SM, writing: AK, CR, critical revisions: all authors, supervision: CR, ARL, RAB, funding: CR, ARL, RAB.

## References

1. Luvira V, Silachamroon U, Piyaphanee W, Lawpoolsri S, Chierakul W, Leaungwutiwong P, et al. Etiologies of Acute Undifferentiated Febrile Illness in Bangkok, Thailand. Am J Trop Med Hyg. 2019;100(3):622–9. Epub 2019/01/11. doi: 10.4269/ajtmh.18-0407. PubMed PMID: 30628565; PubMed Central PMCID: PMCPMC6402898.

2. Sigera C, Rodrigo C, de Silva NL, Weeratunga P, Fernando D, Rajapakse S. Direct costs of managing in-ward dengue patients in Sri Lanka: A prospective study. PloS one. 2021;16(10):e0258388. Epub 2021/10/09. doi: 10.1371/journal.pone.0258388. PubMed PMID: 34624062.

3. Wee LE, Conceicao EP, Sim JX, Aung MK, Oo AM, Yong Y, et al. Dengue and COVID-19: Managing Undifferentiated Febrile Illness during a “Twindemic”. Trop Med Infect Dis. 2022;7(5). Epub 2022/05/28. doi: 10.3390/tropicalmed7050068. PubMed PMID: 35622695; PubMed Central PMCID: PMCPMC9143550.

4. Mørch K, Manoharan A, Chandy S, Singh A, Kuriakose C, Patil S, et al. Clinical features and risk factors for death in acute undifferentiated fever: A prospective observational study in rural community hospitals in six states of India. Transactions of the Royal Society of Tropical Medicine and Hygiene. 2023;117(2):91–101. Epub 2022/09/22. doi: 10.1093/trstmh/trac091. PubMed PMID: 36130240; PubMed Central PMCID: PMCPMC9890314.

5. Wangdi K, Kasturiaratchi K, Nery SV, Lau CL, Gray DJ, Clements ACA. Diversity of infectious aetiologies of acute undifferentiated febrile illnesses in south and Southeast Asia: a systematic review. BMC Infectious Diseases. 2019;19(1):577. doi: 10.1186/s12879-019-4185-y.

6. Susilawati TN, McBride WJ. Undiagnosed undifferentiated fever in Far North Queensland, Australia: a retrospective study. International journal of infectious diseases : IJID : official publication of the International Society for Infectious Diseases. 2014;27:59–64. Epub 2014/09/01. doi: 10.1016/j.ijid.2014.05.022. PubMed PMID: 25173425.

7. Ingarfield SL, Celenza A, Jacobs IG, Riley TV. Outcomes in patients with an emergency department diagnosis of fever of unknown origin. Emergency Medicine Australasia. 2007;19(2):105–12. doi: 10.1111/j.1742-6723.2007.00915.x.

8. Camprubí-Ferrer D, Cobuccio L, Van Den Broucke S, Genton B, Bottieau E, d’Acremont V, et al. Causes of fever in returning travelers: a European multicenter prospective cohort study. Journal of travel medicine. 2022;29(2). Epub 2022/01/19. doi: 10.1093/jtm/taac002. PubMed PMID: 35040473.

9. Lohitharajah J, Malavige GN, Chua AJ, Ng ML, Arambepola C, Chang T. Emergence of human West Nile Virus infection in Sri Lanka. BMC Infect Dis. 2015;15:305. Epub 2015/08/01. doi: 10.1186/s12879-015-1040-7. PubMed PMID: 26227390; PubMed Central PMCID: PMCPMC4521480.

10. Arahirwa V, Tyrlik K, Abernathy H, Cassidy C, Alejo A, Mansour O, et al. Impact of the COVID-19 pandemic on delays in diagnosis and treatment of tick-borne diseases endemic to southeastern USA. Parasites & vectors. 2023;16(1):295. doi: 10.1186/s13071-023-05917-8.

11. Wang S, Xu K, Wang G. Delayed diagnosis of persistent Q fever: a case series from China. BMC infectious diseases. 2024;24(1):591. doi: 10.1186/s12879-024-09484-w.

12. Yufika A, Anwar S, Maulana R, Wahyuniati N, Ramadana RR, Ikram I, et al. Attitude towards Zika among frontline physicians in a dengue-endemic country: A preliminary cross-sectional study in Indonesia. Narra J. 2021;1(1):e32. Epub 2021/04/01. doi: 10.52225/narraj.v1i1.32. PubMed PMID: 38449774; PubMed Central PMCID: PMCPMC10914057.

13. Wang JN, Ling F. Zika Virus Infection and Microcephaly: Evidence for a Causal Link. International journal of environmental research and public health. 2016;13(10). Epub 2016/10/25. doi: 10.3390/ijerph13101031. PubMed PMID: 27775637; PubMed Central PMCID: PMCPMC5086770.

14. Chiu CY, Miller SA. Clinical metagenomics. Nature Reviews Genetics. 2019;20(6):341–55. doi: 10.1038/s41576-019-0113-7.

15. Sigera PC, Amarasekara R, Rodrigo C, Rajapakse S, Weeratunga P, De Silva NL, et al. Risk prediction for severe disease and better diagnostic accuracy in early dengue infection; the Colombo dengue study. BMC infectious diseases. 2019;19(1):680. Epub 2019/08/03. doi: 10.1186/s12879-019-4304-9. PubMed PMID: 31370795; PubMed Central PMCID: PMCPMC6676631.

16. World Health Organization. Dengue guidelines for diagnosis, treatment, prevention and control. New Delhi: WHO; 2009.

17. Sigera PC, Rajapakse S, Weeratunga P, De Silva NL, Gomes L, Malavige GN, et al. Dengue and post-infection fatigue: findings from a prospective cohort-the Colombo Dengue Study. Transactions of the Royal Society of Tropical Medicine and Hygiene. 2020. Epub 2020/10/26. doi: 10.1093/trstmh/traa110. PubMed PMID: 33099653.

18. Zargari Marandi R, Leung P, Sigera C, Murray DD, Weeratunga P, Fernando D, et al. Development of a machine learning model for early prediction of plasma leakage in suspected dengue patients. PLoS neglected tropical diseases. 2023;17(3):e0010758. Epub 2023/03/14. doi: 10.1371/journal.pntd.0010758. PubMed PMID: 36913411; PubMed Central PMCID: PMCPMC10035900.

19. Ministry of Health Sri Lanka. Guidelines on management of dengue fever and dengue haemorrhagic fever in adults. Colombo, Sri Lanka: Ministry of Health Sri Lanka; 2012.

20. Chen S. Ultrafast one-pass FASTQ data preprocessing, quality control, and deduplication using fastp. iMeta. 2023;2(2):e107. doi: 10.1002/imt2.107.

21. Langmead B, Salzberg SL. Fast gapped-read alignment with Bowtie 2. Nature Methods. 2012;9(4):357–9. doi: 10.1038/nmeth.1923.

22. Kim D, Paggi JM, Park C, Bennett C, Salzberg SL. Graph-based genome alignment and genotyping with HISAT2 and HISAT-genotype. Nature Biotechnology. 2019;37(8):907–15. doi: 10.1038/s41587-019-0201-4.

23. Kalantar KL, Carvalho T, de Bourcy CFA, Dimitrov B, Dingle G, Egger R, et al. IDseq—An open source cloud-based pipeline and analysis service for metagenomic pathogen detection and monitoring. GigaScience. 2020;9(10). doi: 10.1093/gigascience/giaa111.

24. Li H. Minimap2: pairwise alignment for nucleotide sequences. Bioinformatics. 2018;34(18):3094–100. doi: 10.1093/bioinformatics/bty191.

25. Buchfink B, Reuter K, Drost H-G. Sensitive protein alignments at tree-of-life scale using DIAMOND. Nature Methods. 2021;18(4):366–8. doi: 10.1038/s41592-021-01101-x.

26. Prjibelski A, Antipov D, Meleshko D, Lapidus A, Korobeynikov A. Using SPAdes De Novo Assembler. Current Protocols in Bioinformatics. 2020;70(1):e102. doi: 10.1002/cpbi.102.

27. Wu TD, Nacu S. Fast and SNP-tolerant detection of complex variants and splicing in short reads. Bioinformatics. 2010;26(7):873–81. doi: 10.1093/bioinformatics/btq057.

28. Ye Y, Choi J-H, Tang H. RAPSearch: a fast protein similarity search tool for short reads. BMC Bioinformatics. 2011;12(1):159. doi: 10.1186/1471-2105-12-159.

29. Edgar RC. MUSCLE: a multiple sequence alignment method with reduced time and space complexity. BMC Bioinformatics. 2004;5(1):113. doi: 10.1186/1471-2105-5-113.

30. Stamatakis A. RAxML version 8: a tool for phylogenetic analysis and post-analysis of large phylogenies. Bioinformatics. 2014;30(9):1312–3. Epub 2014/01/24. doi: 10.1093/bioinformatics/btu033. PubMed PMID: 24451623; PubMed Central PMCID: PMCPMC3998144.

31. Saha S, Ramesh A, Kalantar K, Malaker R, Hasanuzzaman M, Khan LM, et al. Unbiased Metagenomic Sequencing for Pediatric Meningitis in Bangladesh Reveals Neuroinvasive Chikungunya Virus Outbreak and Other Unrealized Pathogens. mBio. 2019;10(6). Epub 2019/12/19. doi: 10.1128/mBio.02877-19. PubMed PMID: 31848287; PubMed Central PMCID: PMCPMC6918088.

32. Sardi SI, Somasekar S, Naccache SN, Bandeira AC, Tauro LB, Campos GS, et al. Coinfections of Zika and Chikungunya Viruses in Bahia, Brazil, Identified by Metagenomic Next-Generation Sequencing. Journal of clinical microbiology. 2016;54(9):2348–53. Epub 2016/07/15. doi: 10.1128/jcm.00877-16. PubMed PMID: 27413190; PubMed Central PMCID: PMCPMC5005514.

33. Souza JVC, Santos HO, Leite AB, Giovanetti M, Bezerra RDS, Carvalho E, et al. Viral Metagenomics for the Identification of Emerging Infections in Clinical Samples with Inconclusive Dengue, Zika, and Chikungunya Viral Amplification. Viruses. 2022;14(9). Epub 2022/09/24. doi: 10.3390/v14091933. PubMed PMID: 36146740; PubMed Central PMCID: PMCPMC9505086.

34. Duong V, Andries AC, Ngan C, Sok T, Richner B, Asgari-Jirhandeh N, et al. Reemergence of Chikungunya virus in Cambodia. Emerging infectious diseases. 2012;18(12):2066–9. Epub 2012/11/23. doi: 10.3201/eid1812.120471. PubMed PMID: 23171736; PubMed Central PMCID: PMCPMC3557864.

35. Quyen NTH, Kien DTH, Rabaa M, Tuan NM, Vi TT, Van Tan L, et al. Chikungunya and Zika Virus Cases Detected against a Backdrop of Endemic Dengue Transmission in Vietnam. The American journal of tropical medicine and hygiene. 2017;97(1):146–50. Epub 2017/07/19. doi: 10.4269/ajtmh.16-0979. PubMed PMID: 28719300; PubMed Central PMCID: PMCPMC5508909.

36. Bohl JA, Lay S, Chea S, Ahyong V, Parker DM, Gallagher S, et al. Discovering disease-causing pathogens in resource-scarce Southeast Asia using a global metagenomic pathogen monitoring system. Proceedings of the National Academy of Sciences. 2022;119(11):e2115285119. doi: 10.1073/pnas.2115285119.

37. Munasinghe DR, Amarasekera PJ, Fernando CF. An epidemic of dengue-like fever in Ceylon (chikungunya--a clinical and haematological study. The Ceylon medical journal. 1966;11(4):129–42. Epub 1966/12/01. PubMed PMID: 5984948.

38. Hermon YE. Virological investigations of Arbovirus infections in Ceylon, with special reference to the recent Chikungunya fever epidemic. The Ceylon medical journal. 1967;12(2):81–92. Epub 1967/06/01. PubMed PMID: 5588665.

39. Obeyesekere I, Hermon Y. Myocarditis and cardiomyopathy after arbovirus infections (dengue and chikungunya fever). British heart journal. 1972;34(8):821–7. Epub 1972/08/01. doi: 10.1136/hrt.34.8.821. PubMed PMID: 4262698; PubMed Central PMCID: PMCPMC486987.

40. Kularatne SA, Gihan MC, Weerasinghe SC, Gunasena S. Concurrent outbreaks of Chikungunya and Dengue fever in Kandy, Sri Lanka, 2006-07: a comparative analysis of clinical and laboratory features. Postgraduate medical journal. 2009;85(1005):342–6. Epub 2009/07/08. doi: 10.1136/pgmj.2007.066746. PubMed PMID: 19581242.

41. Reller ME, Akoroda U, Nagahawatte A, Devasiri V, Kodikaarachchi W, Strouse JJ, et al. Chikungunya as a cause of acute febrile illness in southern Sri Lanka. PloS one. 2013;8(12):e82259. Epub 2013/12/07. doi: 10.1371/journal.pone.0082259. PubMed PMID: 24312651; PubMed Central PMCID: PMCPMC3846738.

42. Hapuarachchi HC, Bandara KB, Sumanadasa SD, Hapugoda MD, Lai YL, Lee KS, et al. Re-emergence of Chikungunya virus in South-east Asia: virological evidence from Sri Lanka and Singapore. The Journal of general virology. 2010;91(Pt 4):1067–76. Epub 2009/12/04. doi: 10.1099/vir.0.015743-0. PubMed PMID: 19955565.

43. Abeygoonawardena H, Wijesinghe N, Navaratne V, Balasuriya A, Nguyen TTN, Moi ML, et al. Serological Evidence of Zika virus Circulation with Dengue and Chikungunya Infections in Sri Lanka from 2017. Journal of global infectious diseases. 2023;15(3):113–20. Epub 2023/10/06. doi: 10.4103/jgid.jgid_195_22. PubMed PMID: 37800085; PubMed Central PMCID: PMCPMC10549900.

44. Ngwe Tun MM, Mutua MM, Inoue S, Takamatsu Y, Kaneko S, Urano T, et al. Molecular and serological evidence of chikungunya virus among dengue suspected patients in Sri Lanka. J Infect Public Health. 2025;18(5):102709. Epub 2025/03/12. doi: 10.1016/j.jiph.2025.102709. PubMed PMID: 40068344.

45. Mavalankar D, Shastri P, Bandyopadhyay T, Parmar J, Ramani KV. Increased mortality rate associated with chikungunya epidemic, Ahmedabad, India. Emerg Infect Dis. 2008;14(3):412–5. Epub 2008/03/08. doi: 10.3201/eid1403.070720. PubMed PMID: 18325255; PubMed Central PMCID: PMCPMC2570824.

46. Rodrigo C, Herath T, Wickramarachchi U, Fernando D, Rajapakse S. Treatment of chikungunya-associated joint pain: a systematic review of controlled clinical trials. Transactions of the Royal Society of Tropical Medicine and Hygiene. 2022;116(10):889–99. Epub 2022/06/07. doi: 10.1093/trstmh/trac045. PubMed PMID: 35666998.

47. Hurley S, Eden JS, Bingham J, Rodriguez M, Neave MJ, Johnson A, et al. Fatal Human Neurologic Infection Caused by Pigeon Avian Paramyxovirus-1, Australia. Emerging infectious diseases. 2023;29(12):2482–7. Epub 2023/11/21. doi: 10.3201/eid2912.230250. PubMed PMID: 37987582; PubMed Central PMCID: PMCPMC10683822.

48. Marinelli T, Masters J, Buckland ME, Lee M, Rawlinson W, Kim KW, et al. Chronic and Neurotropic: A Paradigm-Challenging Case of Dengue Virus Encephalitis in a Patient With Advanced HIV Infection. Clinical infectious diseases : an official publication of the Infectious Diseases Society of America. 2024;79(2):498–501. Epub 2024/02/07. doi: 10.1093/cid/ciae061. PubMed PMID: 38321565.

