## Supplementary Table 1 for "Chikungunya masquerading as dengue infection in Sri Lanka uncovered by metagenomics"

**S1 Table. Viral pathogen reads uncovered with mNGS (only known pathogens).**

| Sample ID | Genomes of pathogens with a positive hit* at a read count filter of > 10 | Number of reads | Genomes of remaining pathogens with at least one positive hit* | Number of reads |
| --- | --- | --- | --- | --- |
| S171 | Human betaherpesvirus 7 | 68 | No additional hits |  |
| S203 | Dengue virus | 1125969 | Cytomegalovirus humanbeta5 | 3 |
|  |  |  | Severe acute respiratory syndrome-related coronavirus | 2 |
|  |  |  | Molluscum contagiosum virus | 2 |
| S197 | Dengue virus | 807 | Severe acute respiratory syndrome-related coronavirus | 2 |
| S162 | Chikungunya virus | 60143 | No additional hits |  |
| S35 | Chikungunya virus | 19395 | No additional hits |  |
| S134 | Chikungunya virus | 2381 | Severe acute respiratory syndrome-related coronavirus | 1 |
| S183 | Chikungunya virus | 114 |  |  |
|  | Severe acute respiratory syndrome-related coronavirus | 11 |  |  |
| S114 | Chikungunya virus | 30 | No additional hits |  |
| S171 | Roseolovirus humanbeta7 | 72 | No additional hits |  |
| S156 |  |  | Cytomegalovirus humanbeta5 | 8 |
|  |  |  | Dengue virus | 2 |
|  |  |  | Severe acute respiratory syndrome-related coronavirus | 1 |
| S64 |  |  | Cytomegalovirus humanbeta5 | 4 |
| S242 |  |  | Betainfluenzavirus influenzae | 8 |
|  |  |  | Severe acute respiratory syndrome-related coronavirus | 1 |
| S11 |  |  | Betainfluenzavirus influenzae | 4 |
| S194 |  |  | Severe acute respiratory syndrome-related coronavirus | 4 |
|  |  |  | Lymphocryptovirus humangamma4 | 2 |
| S137 |  |  | Severe acute respiratory syndrome-related coronavirus | 5 |
|  |  |  | Cytomegalovirus humanbeta5 | 1 |
| S140 |  |  | Severe acute respiratory syndrome-related coronavirus | 2 |
|  |  |  | Cytomegalovirus humanbeta5 | 1 |
| S95 |  |  | Pegivirus hominis | 2 |
| S146 |  |  | Severe acute respiratory syndrome-related coronavirus | 1 |
| S200 |  |  | Severe acute respiratory syndrome-related coronavirus | 1 |
| S154 |  |  | Betapolyomavirus secuhominis | 4 |
| S34 |  |  | Human immunodeficiency virus | 4 |
|  |  |  | Cytomegalovirus humanbeta5 | 1 |
| S127 |  |  | Betapolyomavirus quartihominis | 4 |
|  |  |  | Severe acute respiratory syndrome-related coronavirus | 2 |
| S235 |  |  | Alphainfluenzavirus influenzae | 8 |
|  |  |  | Severe acute respiratory syndrome-related coronavirus | 1 |
| S98 | Alphapolyomavirus quintihominis | 32 | No additional hits |  |
| S250 |  |  | Severe acute respiratory syndrome-related coronavirus | 4 |
|  |  |  | Alphapolyomavirus quintihominis | 4 |
| S236 | Dengue virus | 147733 | No additional hits |  |
| S158 | Dengue virus | 2754 | No additional hits |  |
| S226 | Dengue virus | 460 | Severe acute respiratory syndrome-related coronavirus | 1 |
| S51 |  |  | Lymphocryptovirus humangamma4 | 6 |
|  |  |  | Cytomegalovirus humanbeta5 | 2 |
|  |  |  | Morbillivirus hominis | 1 |
| S164 |  |  | Human betaherpesvirus 6 | 4 |
|  |  |  | Cytomegalovirus humanbeta5 | 2 |
| S247 |  |  | Dengue virus | 6 |
| S160 |  |  | Dengue virus | 2 |
| S66 |  |  | Cytomegalovirus humanbeta5 | 2 |
| S84 |  |  | Cytomegalovirus humanbeta5 | 2 |
| S70 |  |  | Cytomegalovirus humanbeta5 | 1 |
| S16 |  |  | Cytomegalovirus humanbeta5 | 1 |
| S166 | Chikungunya virus | 1630 | Cytomegalovirus humanbeta5 | 1 |
| S79 |  |  | Molluscum contagiosum virus | 1 |
| S77 |  |  | Human betaherpesvirus 6 | 2 |
|  |  |  | Roseolovirus humanbeta6b | 8 |
| S145 |  |  | Roseolovirus humanbeta7 | 2 |
| S105 |  |  | Chikungunya virus | 4 |
| S112 | Chikungunya virus | 25401 | No additional hits |  |

*Each “hit” is a 150-300bp length single Illumina read out of millions for that sample with a number of mismatches to reference genome and hence unlikely to represent a true infection.
