## Supplementary Table 2 for "Chikungunya masquerading as dengue infection in Sri Lanka uncovered by metagenomics"

**S2 Table**. **Pathogenic bacterial reads uncovered with mNGS (only known pathogens).**

| Sample ID | Genomes of pathogens with a positive hit* at a read count filter of >100 rPM | Number of reads | Genomes of pathogens with a positive hit* at a read count filter of 10 < rPM < 100 | Number of reads |
| --- | --- | --- | --- | --- |
| S134 | Pseudomonas putida | 239903 | Aeromonas caviae | 5798 |
|  | Acinetobacter baumannii | 10841 | Elizabethkingia anophelis | 5290 |
|  | Escherichia coli | 7029 | Burkholderia cepacia | 5165 |
|  | Klebsiella pneumoniae | 6900 | Aeromonas veronii | 4305 |
|  | Burkholderia cenocepacia | 15488 | Stenotrophomonas maltophilia | 2999 |
|  | Enterobacter cloacae | 16519 | Citrobacter freundii | 2808 |
|  |  |  | Pseudomonas aeruginosa | 2796 |
|  |  |  | Yersinia pestis | 1849 |
|  |  |  | Enterococcus faecalis | 1127 |
|  |  |  | Aeromonas hydrophila | 1052 |
|  |  |  | Staphylococcus epidermidis | 850 |
| S137 | Pseudomonas putida | 188452 | Aeromonas caviae | 6037 |
|  | Acinetobacter baumannii | 165722 | Escherichia coli | 4390 |
|  | Burkholderia cenocepacia | 34716 | Salmonella enterica | 1027 |
|  | Enterobacter cloacae | 26757 | Pseudomonas fluorescens | 2618 |
|  |  |  | Burkholderia cepacia | 5997 |
|  |  |  | Klebsiella pneumoniae | 3507 |
|  |  |  | Stenotrophomonas maltophilia | 1693 |
|  |  |  | Citrobacter freundii | 1579 |
|  |  |  | Pseudomonas aeruginosa | 6352 |
|  |  |  | Streptococcus mitis | 2550 |
|  |  |  | Haemophilus parainfluenzae | 1725 |
|  |  |  | Streptococcus oralis | 2181 |
|  |  |  | Klebsiella aerogenes | 4969 |
|  |  |  | Staphylococcus epidermidis | 1149 |
| S156 | Stenotrophomonas maltophilia | 16053 | Burkholderia cepacia | 7412 |
|  | Pseudomonas putida | 44466 | Enterobacter cloacae | 2404 |
|  | Acinetobacter baumannii | 45614 | Burkholderia cenocepacia | 7946 |
|  |  |  | Pseudomonas aeruginosa | 8067 |
|  |  |  | Burkholderia pseudomallei | 1505 |
| S140 | Pseudomonas aeruginosa | 74408 | Aeromonas caviae | 11328 |
|  | Stenotrophomonas maltophilia | 55761 | Escherichia coli | 2999 |
|  | Pseudomonas putida | 23592 | Agrobacterium tumefaciens | 12967 |
|  | Acinetobacter baumannii | 21418 | Burkholderia cepacia | 14154 |
|  | Achromobacter xylosoxidans | 25725 | Klebsiella pneumoniae | 2351 |
|  | Burkholderia cenocepacia | 46493 | Citrobacter freundii | 1832 |
|  | Enterobacter cloacae | 32361 | Ralstonia pickettii | 4690 |
|  |  |  | Staphylococcus epidermidis | 1743 |
| S110 | Pseudomonas aeruginosa | 9182 | Pseudomonas putida | 3660 |
|  | Stenotrophomonas maltophilia | 14585 | Escherichia coli | 2137 |
|  | Acinetobacter baumannii | 7089 | Salmonella enterica | 1379 |
|  |  |  | Pseudomonas fluorescens | 2215 |
|  |  |  | Achromobacter xylosoxidans | 963 |
|  |  |  | Burkholderia cepacia | 4060 |
|  |  |  | Enterobacter cloacae | 1382 |
|  |  |  | Klebsiella pneumoniae | 4579 |
|  |  |  | Burkholderia cenocepacia | 2433 |
|  |  |  | Methylorubrum extorquens | 5548 |
|  |  |  | Burkholderia pseudomallei | 1686 |
|  |  |  | Ralstonia pickettii | 1325 |
|  |  |  | Enterococcus faecalis | 3761 |
| S127 | Pseudomonas aeruginosa | 27120 | Elizabethkingia anophelis | 2903 |
|  | Enterobacter cloacae | 20527 | Pseudomonas putida | 11160 |
|  |  |  | Pseudomonas fluorescens | 1768 |
| S27 |  |  | Pseudomonas putida | 1870 |
|  |  |  | Enterobacter cloacae | 4615 |
|  |  |  | Stenotrophomonas maltophilia | 1696 |
|  |  |  | Pseudomonas aeruginosa | 10196 |
| S98 |  |  | Agrobacterium tumefaciens | 5711 |
|  |  |  | Elizabethkingia anophelis | 3942 |
|  |  |  | Stenotrophomonas maltophilia | 2846 |
|  |  |  | Acinetobacter baumannii | 2835 |
| S34 |  |  | Stenotrophomonas maltophilia | 6152 |
|  |  |  | Enterobacter cloacae | 3404 |
|  |  |  | Acinetobacter baumannii | 2573 |
|  |  |  | Escherichia coli | 2170 |
|  |  |  | Staphylococcus epidermidis | 2134 |
|  |  |  | Citrobacter freundii | 2073 |
|  |  |  | Salmonella enterica | 1644 |
| S64 |  |  | Pseudomonas aeruginosa | 1726 |
| S11 |  |  | Pseudomonas aeruginosa | 1715 |
| S114 |  |  | Staphylococcus epidermidis | 1868 |
| S250 |  |  | Stenotrophomonas maltophilia | 3957 |
|  |  |  | Staphylococcus epidermidis | 2984 |
|  |  |  | Escherichia coli | 2425 |
|  |  |  | Staphylococcus aureus | 1673 |
|  |  |  | Acinetobacter baumannii | 1452 |
| S194 |  |  | Escherichia coli | 3816 |
|  |  |  | Pseudomonas putida | 2074 |
| S242 |  |  | Acinetobacter baumannii | 1861 |
| S35 |  |  | Pseudomonas aeruginosa | 7786 |
|  |  |  | Pseudomonas putida | 3941 |
|  |  |  | Methylorubrum extorquens | 1569 |
| S200 |  |  | Aeromonas caviae | 3228 |
|  |  |  | Pseudomonas aeruginosa | 2627 |
| S171 | Pseudomonas aeruginosa | 18883 | Burkholderia pseudomallei | 601 |
|  | Stenotrophomonas maltophilia | 7982 | Aeromonas hydrophila | 524 |
|  | Pseudomonas putida | 6622 | Aeromonas veronii | 450 |
|  | Acinetobacter baumannii | 5324 | Klebsiella pneumoniae | 419 |
|  | Agrobacterium tumefaciens | 3139 | Methylorubrum extorquens | 338 |
|  | Aeromonas caviae | 16434 | Burkholderia cenocepacia | 334 |
|  | Citrobacter freundii | 1115 | Achromobacter xylosoxidans | 317 |
|  |  |  | Escherichia coli | 314 |
|  |  |  | Salmonella enterica | 303 |
| S95 | Klebsiella pneumoniae | 27965 | Pseudomonas aeruginosa | 4783 |
|  |  |  | Agrobacterium tumefaciens | 1885 |
|  |  |  | Stenotrophomonas maltophilia | 1879 |
|  |  |  | Achromobacter xylosoxidans | 1575 |
| S170 |  |  | Stenotrophomonas maltophilia | 3707 |
|  |  |  | Ralstonia pickettii | 1959 |
|  |  |  | Agrobacterium tumefaciens | 1906 |
|  |  |  | Achromobacter xylosoxidans | 1594 |
|  |  |  | Pseudomonas putida | 1512 |
| S57 |  |  | Pseudomonas aeruginosa | 3549 |
|  |  |  | Stenotrophomonas maltophilia | 2933 |
|  |  |  | Bordetella bronchiseptica | 2021 |
|  |  |  | Achromobacter xylosoxidans | 1603 |
| S183 |  |  | Haemophilus parainfluenzae | 8692 |
|  |  |  | Pseudomonas aeruginosa | 5814 |
|  |  |  | Escherichia coli | 2186 |
|  |  |  | Pseudomonas putida | 1991 |
|  |  |  | Stenotrophomonas maltophilia | 1519 |
|  |  |  | Aggregatibacter actinomycetemcomitans | 1434 |
|  |  |  | Achromobacter xylosoxidans | 1424 |
|  |  |  | Acinetobacter baumannii | 1401 |
|  |  |  | Agrobacterium tumefaciens | 1318 |
| S154 |  |  | Staphylococcus epidermidis | 1568 |
| S203 | Staphylococcus epidermidis | 31280 | Pseudomonas putida | 6262 |
|  |  |  | Streptococcus mitis | 2110 |
|  |  |  | Edwardsiella tarda | 1849 |
|  |  |  | Escherichia coli | 1689 |
|  |  |  | Eikenella corrodens | 1653 |
|  |  |  | Acinetobacter baumannii | 1524 |
|  |  |  | Stenotrophomonas maltophilia | 1485 |
|  |  |  | Anaerococcus prevotii | 1430 |
| S51 | Pseudomonas putida | 24269 | Salmonella enterica | 11129 |
|  | Stenotrophomonas maltophilia | 106644 | Pseudomonas aeruginosa | 7198 |
|  |  |  | Pseudomonas fluorescens | 6809 |
|  |  |  | Klebsiella pneumoniae | 6189 |
|  |  |  | Acinetobacter baumannii | 4137 |
|  |  |  | Citrobacter freundii | 4092 |
|  |  |  | Agrobacterium tumefaciens | 3249 |
|  |  |  | Escherichia coli | 3036 |
|  |  |  | Staphylococcus epidermidis | 2429 |
|  |  |  | Burkholderia cenocepacia | 2202 |
| S43 | Pseudomonas aeruginosa | 32585 | Stenotrophomonas maltophilia | 4983 |
|  | Pseudomonas putida | 29098 | Agrobacterium tumefaciens | 4521 |
|  | Methylorubrum extorquens | 30117 | Serratia marcescens | 3068 |
|  |  |  | Pseudomonas fluorescens | 2158 |
|  |  |  | Aeromonas caviae | 2045 |
| S105 | Pseudomonas putida | 131729 | Elizabethkingia anophelis | 1782 |
|  |  |  | Agrobacterium tumefaciens | 3486 |
|  |  |  | Pseudomonas aeruginosa | 3202 |
| S66 |  |  | Elizabethkingia anophelis | 1793 |
|  |  |  | Salmonella enterica | 1266 |
|  |  |  | Pseudomonas aeruginosa | 756 |
|  |  |  | Acinetobacter baumannii | 692 |
|  |  |  | Staphylococcus epidermidis | 604 |
| S66 |  |  | Pseudomonas aeruginosa | 7126 |
|  |  |  | Stenotrophomonas maltophilia | 6242 |
| S104 | Pseudomonas putida | 111002 | Staphylococcus epidermidis | 3460 |
|  | Agrobacterium tumefaciens | 9971 | Agrobacterium tumefaciens | 2338 |
|  |  |  | Acinetobacter baumannii | 1940 |
|  |  |  | Pseudomonas aeruginosa | 1138 |
|  |  |  | Elizabethkingia anophelis | 1033 |
| S104 | Pseudomonas putida | 21074 | Pseudomonas aeruginosa | 2833 |
|  |  |  | Acinetobacter baumannii | 2071 |
|  |  |  | Staphylococcus epidermidis | 1939 |
|  |  |  | Methylorubrum extorquens | 1886 |
|  |  |  | Escherichia coli | 748 |
| S61 |  |  | Pseudomonas aeruginosa | 3203 |
|  |  |  | Agrobacterium tumefaciens | 1423 |
|  |  |  | Elizabethkingia anophelis | 1316 |
|  |  |  | Stenotrophomonas maltophilia | 1277 |
| S85 |  |  | Pseudomonas aeruginosa | 1088 |
| S77 | Acinetobacter baumannii | 18786 | Acinetobacter baumannii | 4391 |
|  | Ralstonia picketti | 12782 | Enterobacter cloacae | 2247 |
|  | Bacillus cereus | 32913 | Elizabethkingia anophelis | 1198 |
|  |  |  | Pseudomonas aeruginosa | 1020 |
| S112 |  |  | Elizabethkingia anophelis | 3308 |
|  |  |  | Pseudomonas aeruginosa | 1721 |
| S52 |  |  | Pseudomonas aeruginosa | 3669 |
|  |  |  | Elizabethkingia anophelis | 903 |
| S79 |  |  | Enterobacter cloacae | 1586 |
| S70 | Aeromonas caviae | 21444 | Elizabethkingia anophelis | 4335 |
|  |  |  | Enterobacter cloacae | 1294 |
|  |  |  | Acinetobacter baumannii | 1218 |
|  |  |  | Staphylococcus epidermidis | 572 |
| S167 |  |  | Actinomyces israelii | 11 |
|  |  |  | Elizabethkingia anophelis | 10 |
|  |  |  | Staphylococcus aureus | 9 |
|  |  |  | Streptococcus sanguinis | 8 |
|  |  |  | Staphylococcus saprophyticus | 7 |
|  |  |  | Streptococcus pneumoniae | 6 |
|  |  |  | Nocardia nova | 4 |
|  |  |  | Vibrio parahaemolyticus | 4 |
|  |  |  | Enterobacter cloacae | 3 |
|  |  |  | Corynebacterium diphtheriae | 3 |
| S145 |  |  | Streptococcus oralis | 2146 |
|  |  |  | Streptococcus pneumoniae | 1177 |
|  |  |  | Streptococcus mitis | 873 |
| S122 |  |  | Acinetobacter baumannii | 3878 |
