## Supplementary Table 3 for "Chikungunya masquerading as dengue infection in Sri Lanka uncovered by metagenomics"

S3 Table. **An age and gender matched 1:2 comparison between confirmed chikungunya and dengue cases (N=15)**

| **Parameter** | **Dengue infection (n = 10)** | | **Chikungunya infection (n = 5)** | | **P-value** |
| --- | --- | --- | --- | --- | --- |
|  | **Median** | **Inter-quartile range** | **Median** | **Inter-quartile range** |  |
| ***Biochemical* (serum)*** |  |  |  |  |  |
| Sodium (mmol/l) | 135 | 133 - 139 | 135 | 133.5 – 135.5 | 0.606 |
| Potassium (mmol/l) | 3.3 | 3 – 3.4 | 4.2 | 3.6 – 6.4 | 0.012 |
| ALT (IU/l) | 33 | 22.8 – 60.3 | 46 | 6 - 77 | 0.733 |
| AST (IU/l) | 33 | 28 – 60.5 | 29.5 | 18.8 – 105.5 | 0.539 |
| CRP (mg/l) | 7 | 6 – 11.7 | 18 | 6 – 77.5 | 0.371 |
| Creatinine (µmol/l) | 85 | 77.2 – 102.8 | 94 | 70 - 135 | 0.594 |
| ***Haematological***** |  |  |  |  |  |
| Total white cell count (*10^3^/µl) | 2.9 | 2.3 – 3.8 | 3.9 | 2.6 – 5.8 | 0.310 |
| Neutrophils (*10^3^/µl) | 1.4 | 1 – 1.8 | 1.7 | 1 – 2.8 | 0.44 |
| Lymphocytes (*10^3^/µl) | 1 | 0.6 – 1.8 | 1.5 | 0.8 – 2.8 | 0.513 |
| Platelets (*10^3^/µl) | 101.5 | 75.2 – 124.7 | 112 | 83 - 153 | 0.513 |
| Haemoglobin (g/dl) | 13.9 | 13.1 – 14.7 | 12.8 | 7.9 – 13.5 | 0.055 |
| Haematocrit (%) | 43.5 | 39.9 – 44.6 | 37.1 | 23.4 – 39.1^#^ | 0.003 |
| *Symptom score**** | 3 | 1.75 - 4 | 4 | 3.5 - 4 | 0.254 |
| *Hospital stay (days)* | 3.2 | 2.2 – 4.4 | 2.6 | 2.2 -4.4 | 0.859 |

AST – Aspartate aminotransferase, ALT – Alanine aminotransferase, CRP – C reactive protein, NS – Not statistically significant with Mann-Whitney U test (Bonferroni adjusted p-value of >0.004). * First reported value within the first 96 hours of fever, **value reported on day 3 of fever and when multiple tests were done per day, the lowest WBC, neutrophil, lymphocyte, haemoglobin, platelet counts and highest haematocrit value was considered. *** calculated as sum of the presence (1) or absence (0) of abdominal pain, arthralgia, gum bleeding, other bleeding manifestations, cough, chills or rigors, diarrhoea, dyspnoea, headache, retro-orbital pain, myalgia and nausea/vomiting on admission.
